# A 56-Day Single-Arm Exploratory Study of NatureU® Mind Care BeautyU Caps on Crow’s-Feet Wrinkle Count, Skin Hydration, and Related Skin-Aging Parameters in Adult Women

**DOI:** 10.64898/2026.05.07.26351904

**Authors:** Lorry Luo, Luke Law, Naining Zhang

## Abstract

**Background:** Skin aging is multifactorial, and finished multi-ingredient oral beauty supplements require dedicated clinical evaluation because their effects cannot be inferred from individual ingredient data alone.

**Objective:** To explore, in a 56-day single-arm open-label study, whether daily oral intake of NatureU® Mind Care BeautyU Caps is associated with within-participant changes in crow’s-feet wrinkle count (primary endpoint), stratum corneum hydration (secondary endpoint), and additional exploratory skin-aging parameters in adult women.

**Methods:** A single-center, open-label, single-arm exploratory study enrolled 33 healthy women aged 36–56 years, eligible only if baseline cheek hydration was below 60 corneometer units (C.U.); 31 completed the protocol. Participants took one capsule orally once daily for 56 consecutive days. Assessments at D0, D28 and D56 used PRIMOS CR, Corneometer CM 825, Cutometer MPA580, Glossymeter, Colorimeter CL400, Mexameter MX18, VISIA CR, DermaScan and a structured self-assessment.

**Results:** PRIMOS CR crow’s-feet wrinkle count fell from 965.3 ± 333.7 at D0 to 514.2 ± 171.0 at D56 (−46.73%; Wilcoxon P = 1.2 × 10□□), and Corneometer hydration rose from 44.3 ± 7.8 to 70.3 ± 9.9 (+58.57%; P = 1.2 × 10□□). Both changed in all 31 completers. Average wrinkle depth changed only non-significantly (−4.81%; P = 0.058), decreasing in 19 participants, increasing in 11 and unchanged in one. Individual-participant data obtained after version 1 show that the reduction in wrinkle count was almost exactly proportional across a 5.9-fold range of baseline values (D56/D0 ratio 0.535 ± 0.041; coefficient of variation 7.7%, lower than any other endpoint), was uncorrelated with baseline severity (r = −0.15, P = 0.42), was complete by D28 (D56/D28 ratio 0.990 ± 0.096), and was uncorrelated with the change in wrinkle depth (r = +0.30, P = 0.096) or hydration (r = +0.23, P = 0.21). No adverse reactions were reported.

**Conclusion:** Daily intake over 56 days was associated with an increase in stratum corneum hydration that cannot be accounted for by the low-hydration entry criterion alone. A reduction in crow’s-feet wrinkle count was also recorded, but its uniformity across participants, its independence of baseline severity, its completion at the first follow-up visit, and the absence of the analysis parameters needed to reproduce it mean that the authors do not interpret it as evidence of a biological anti-wrinkle effect. This conclusion supersedes that of version 1. Findings are hypothesis-generating; randomized, placebo-controlled trials are required.

**Version note — what changed and why:** This version revises version 1 following peer review of the same study. Readers should not rely on version 1. Six substantive changes have been made.

1. **The interpretation of the primary endpoint has changed** After version 1 was posted, the authors obtained the individual-participant dataset from the contract testing organization. It shows that the reduction in crow’s-feet wrinkle count occurred in all 31 completers, was almost exactly proportional across a 5.9-fold range of baseline values, was uncorrelated with baseline severity, was complete at day 28 with no further progression, and was uncorrelated with the concurrent change in wrinkle depth or hydration. The acquisition and analysis parameters needed to exclude a measurement explanation — the PRIMOS segmentation threshold, the region-of-interest definition and whether images were spatially registered across timepoints — were requested from the contract testing organization and were not provided. **The authors therefore no longer interpret the reduction in wrinkle count as evidence of a biological anti-wrinkle effect**. Version 1 did interpret it that way; that interpretation is withdrawn.
2. **The description of the ethics committee has been corrected** Version 1 described the review body as a “dedicated ethics oversight body”. It is the internal ethics committee of the contract testing organization that conducted the study under commission from, and for payment by, the sponsor. The corrected Ethics statement below says so explicitly.
3. **The trial registration statement has been corrected** Version 1 contained an internal inconsistency: the body text stated that the study “was not prospectively registered”, while the submission metadata gave a registration identifier and answered the registration declaration affirmatively without the explanation that medRxiv requires for retrospectively registered studies. The corrected statement below gives the registration date and the interval.
4. **The conflict-of-interest statement has been made specific** Version 1 stated only that authors affiliated with the sponsor “declare those relationships in accordance with the journal’s disclosure requirements”, which disclosed nothing. The corrected statement below names the relationships.
5. **An appendix has been removed** Version 1 reproduced, in full, an administrative document titled “Ethics Review and Regulatory Classification Report”, including a regulatory classification of the product under Hong Kong law. That document is not study content, its regulatory classification is not a finding of this study, and its description of the ethics committee is superseded by item 2 above. It has been removed. It remains available in version 1 for anyone who wishes to consult it.
6. **The author byline has been corrected** Version 1 listed four authors in the manuscript byline. One of those names, Miexin Yang, was included in the manuscript file in error: that person did not contribute to the work reported here and was never registered as an author in the submission record, which has listed three authors throughout. The byline in this version therefore lists the three authors of record — Lorry Luo, Luke Law and Naining Zhang — and now matches the submission metadata. No author has been added, and no author who contributed to the work has been removed.

The affiliation of Naining Zhang has been updated from “CTI AiPu Medical Laboratory (Shanghai) Co., Ltd.” to “Centre Testing International Pinbiao (Shanghai) Co., Ltd.”. These are two English renderings of the same legal entity; the latter is the name that appears on the ethics approval and is used throughout this version.

**Figures and tables:** Version 1 contained two figures and no data tables. This version retains both (Figure 1 has been redrawn so that the three columns are no longer joined by causal arrows, because no such link was measured) and adds Figure 3 and Tables 1–5, which present the underlying summary data in full. Two panels of photographic material that accompanied version 1’s supplementary documentation — a commercial wrinkle-grading reference scale and example instrument output images — have not been carried over, since they are photographs of skin and are not required to follow the analysis.

Three numerical corrections have also been made. Wrinkle depth decreased in 19 of 31 participants, increased in 11 and was unchanged in one; version 1 and an earlier draft of this version reported “12” increases, which omitted the unchanged participant. Percentage changes are now computed throughout from unrounded means, which changes the reported day-28 hydration increase from +65.01% to +64.77%; the testing report computed these percentages from means rounded to one decimal place. The correlation statistics now state the quantity on which each was computed.

The study data are unchanged. No participant data, measurement or summary statistic has been altered; the recomputed values reported below were derived by the authors from the individual-participant dataset.

## 1. Introduction

Skin aging reflects intrinsic chronological aging and extrinsic environmental exposures (ultraviolet radiation, pollution, oxidative stress and chronic low-grade inflammation), producing dryness, reduced elasticity, wrinkles, pigmentary unevenness, reduced radiance and dermal remodeling (Russell-Goldman and Murphy, 2020; D’Arino et al., 2022; Park et al., 2023). Mechanistic drivers include oxidative damage, mitochondrial dysfunction, cellular senescence, inflammatory signaling and matrix metalloproteinase–mediated degradation of collagen and elastin (Liu et al., 2023a; Hu et al., 2023).

**Table 1.** Study design, endpoint hierarchy, participant disposition and tolerability.

| Item | Value |
| --- | --- |
| Study design | Single-centre, open-label, single-arm exploratory clinical study |
| Intervention | NatureU® Mind Care BeautyU Caps, one capsule orally once daily |
| Study duration | 56 days |
| Assessment timepoints | Baseline (D0), day 28 (D28), day 56 (D56) |
| Primary endpoint | D56 change from baseline in PRIMOS CR crow's-feet wrinkle count |
| Secondary endpoint | D56 change from baseline in Corneometer CM 825 stratum corneum hydration |
| Exploratory endpoints | Other PRIMOS CR wrinkle parameters (length, volume, depth, area, area ratio); Cutometer MPA580 elasticity (R2, F3/F4, F4); Glossometer and VISIA CR gloss/radiance; Colorimeter CL400 and VISIA CR ITA°; Mexameter MX18 melanin index; VISIA CR spot area ratio, spot optical density, pore score, pore feature count; DermaScan dermal thickness and density; participant-reported outcomes |
| Analysis set | Completer analysis ($n = 31$); no imputation |
| Power calculation | Not performed (exploratory, signal-generating evaluation) |
| Multiplicity correction | None applied; P values are nominal |
| Target participant profile | Healthy adult women aged 36–56 years with cheek hydration below 60 C.U., visible crow's-feet wrinkles, facial laxity or reduced radiance, and at least one visible facial spot meeting prespecified criteria |
| Candidates screened | 56 |
| Participants enrolled | 33 |
| Participants completing the intervention | 31 (93.9%) |
| Withdrawals | 2 (one lost to follow-up at D56 after contracting influenza A; one withdrew on day 4 because of illness) |
| Withdrawals attributed to adverse reactions or intolerance | 0 |
| Self-reported adverse reactions during the 56-day intervention | None reported among the 31 evaluable participants |
| Overall tolerability | The product was well tolerated under the conditions of this exploratory 56-day study; this conclusion remains limited by the open-label design, the sample size and the study duration |

**Table 2.**
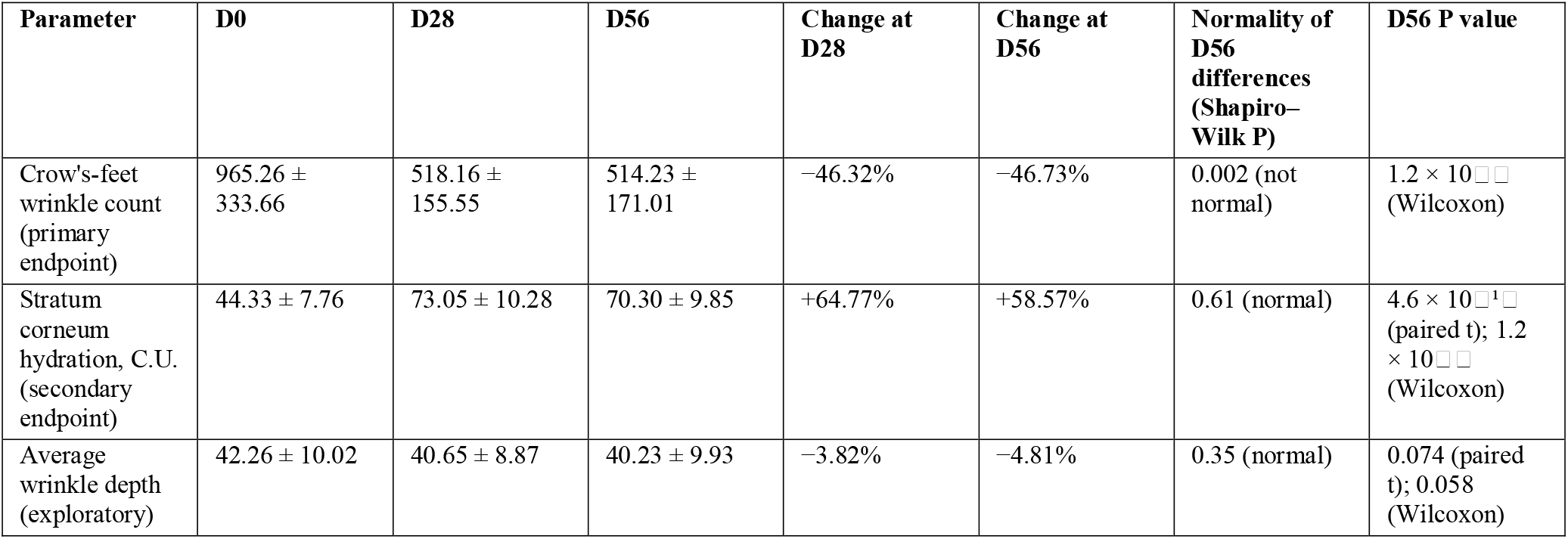
Primary endpoint, secondary endpoint and average wrinkle depth, recomputed by the authors from the individual-participant dataset (n = 31 completers). Values are mean ± standard deviation. Percentage changes are computed from unrounded means; the original testing report computed them from means rounded to one decimal place and therefore reports +65.01% and +58.69% for hydration at D28 and D56, and −4.80% for wrinkle depth at D56.

**Table 3.** PRIMOS CR wrinkle-related outcomes (n = 31 completers). Values are mean ± standard deviation as reported by the contract testing organization. P values are nominal and unadjusted for multiplicity.

| Parameter | D0 | D28 | D56 | Change at D28 | Change at D56 | D28 P value | D56 P value |
| --- | --- | --- | --- | --- | --- | --- | --- |
| Wrinkle count (primary endpoint) | 965 ± 334 | 518 ± 156 | 514 ± 171 | -46.32% | -46.73% | < 0.001 | < 0.001 |
| Wrinkle length, mm | 303.97 ± 83.08 | 234.74 ± 66.11 | 232.00 ± 60.90 | -22.78% | -23.68% | < 0.001 | < 0.001 |
| Wrinkle volume | 4.18 ± 1.45 | 3.31 ± 1.00 | 3.29 ± 1.07 | -20.81% | -21.29% | < 0.001 | < 0.001 |
| Wrinkle depth | 42.26 ± 10.02 | 40.65 ± 8.87 | 40.23 ± 9.93 | -3.82% | -4.81% | 0.310 | 0.058 |
| Wrinkle area | 99.13 ± 27.08 | 82.51 ± 23.58 | 82.15 ± 22.33 | -16.77% | -17.13% | < 0.001 | < 0.001 |
| Wrinkle area ratio | 19.34% ± 0.17% | 16.07% ± 0.78% | 16.04% ± 0.68% | -16.91% | -17.06% | < 0.001 | < 0.001 |

**Table 4.**
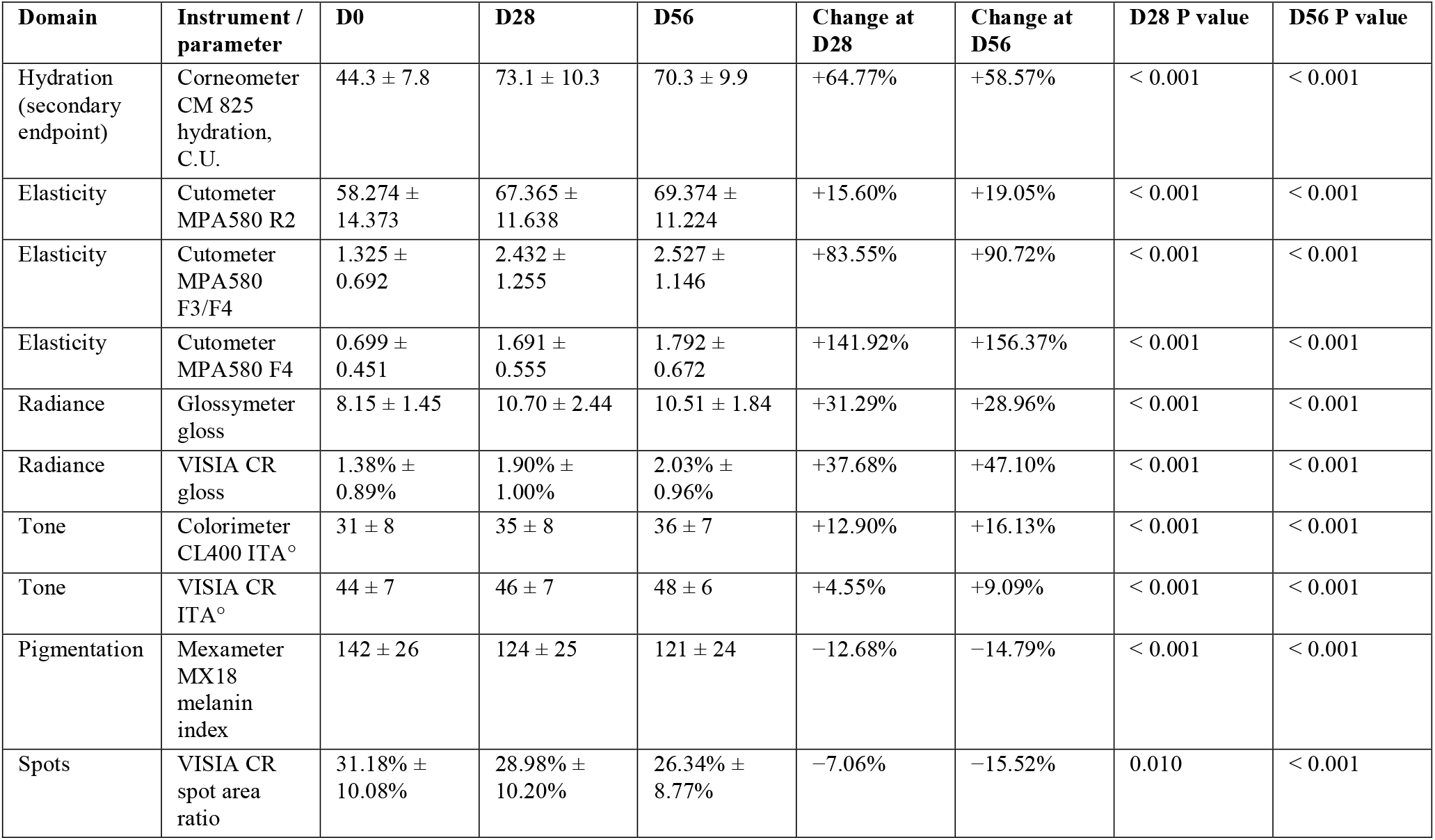
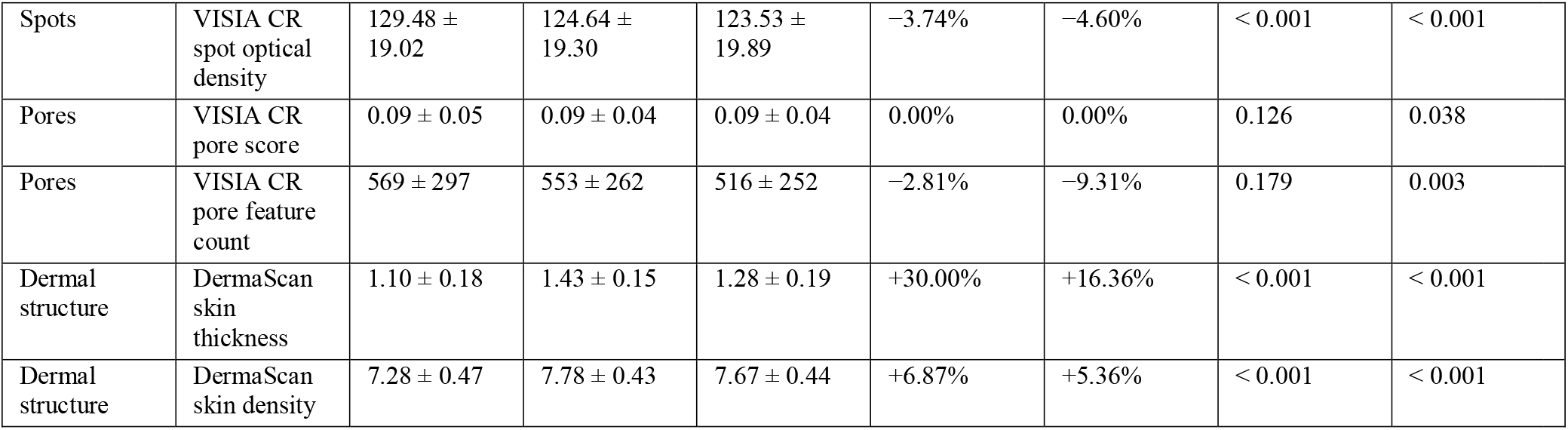
Non-wrinkle instrumental outcomes (n = 31 completers). Values are mean ± standard deviation as reported by the contract testing organization. P values are nominal and unadjusted for multiplicity. All parameters other than Corneometer hydration at D56 are exploratory.

| Domain | Instrument / parameter | D0 | D28 | D56 | Change at D28 | Change at D56 | D28 P value | D56 P value |
| --- | --- | --- | --- | --- | --- | --- | --- | --- |
| Hydration (secondary endpoint) | Corneometer CM 825 hydration, C.U. | 44.3 ± 7.8 | 73.1 ± 10.3 | 70.3 ± 9.9 | +64.77% | +58.57% | < 0.001 | < 0.001 |
| Elasticity | Cutometer MPA580 R2 | 58.274 ± 14.373 | 67.365 ± 11.638 | 69.374 ± 11.224 | +15.60% | +19.05% | < 0.001 | < 0.001 |
| Elasticity | Cutometer MPA580 F3/F4 | 1.325 ± 0.692 | 2.432 ± 1.255 | 2.527 ± 1.146 | +83.55% | +90.72% | < 0.001 | < 0.001 |
| Elasticity | Cutometer MPA580 F4 | 0.699 ± 0.451 | 1.691 ± 0.555 | 1.792 ± 0.672 | +141.92% | +156.37% | < 0.001 | < 0.001 |
| Radiance | Glossometer gloss | 8.15 ± 1.45 | 10.70 ± 2.44 | 10.51 ± 1.84 | +31.29% | +28.96% | < 0.001 | < 0.001 |
| Radiance | VISIA CR gloss | 1.38% ± 0.89% | 1.90% ± 1.00% | 2.03% ± 0.96% | +37.68% | +47.10% | < 0.001 | < 0.001 |
| Tone | Colorimeter CL400 ITA° | 31 ± 8 | 35 ± 8 | 36 ± 7 | +12.90% | +16.13% | < 0.001 | < 0.001 |
| Tone | VISIA CR ITA° | 44 ± 7 | 46 ± 7 | 48 ± 6 | +4.55% | +9.09% | < 0.001 | < 0.001 |
| Pigmentation | Mexameter MX18 melanin index | 142 ± 26 | 124 ± 25 | 121 ± 24 | -12.68% | -14.79% | < 0.001 | < 0.001 |
| Spots | VISIA CR spot area ratio | 31.18% ± 10.08% | 28.98% ± 10.20% | 26.34% ± 8.77% | -7.06% | -15.52% | 0.010 | < 0.001 |
| Spots | VISIA CR spot optical density | 129.48 ± 19.02 | 124.64 ± 19.30 | 123.53 ± 19.89 | -3.74% | -4.60% | < 0.001 | < 0.001 |
| Pores | VISIA CR pore score | 0.09 ± 0.05 | 0.09 ± 0.04 | 0.09 ± 0.04 | 0.00% | 0.00% | 0.126 | 0.038 |
| Pores | VISIA CR pore feature count | 569 ± 297 | 553 ± 262 | 516 ± 252 | -2.81% | -9.31% | 0.179 | 0.003 |
| Dermal structure | DermaScan skin thickness | 1.10 ± 0.18 | 1.43 ± 0.15 | 1.28 ± 0.19 | +30.00% | +16.36% | < 0.001 | < 0.001 |
| Dermal structure | DermaScan skin density | 7.28 ± 0.47 | 7.78 ± 0.43 | 7.67 ± 0.44 | +6.87% | +5.36% | < 0.001 | < 0.001 |

**Table 5.** Participant-reported recognition rates for questionnaire items 1–19 (n = 31 completers), with P values recomputed by the authors using a two-sided exact binomial test against the prespecified 60% benchmark. Recognition rate is the proportion of participants selecting an agreement-level response. The D28 P values differ from those in the original testing report for the reason given in Section 2.6. Item wording is given in Appendix Table A2.

| Item | D28 recognition rate | D28 P value (recomputed) | D56 recognition rate | D56 P value (recomputed) |
| --- | --- | --- | --- | --- |
| 1 | 87.10% | 0.002 | 96.77% | < 0.001 |
| 2 | 96.77% | < 0.001 | 100.00% | < 0.001 |
| 3 | 96.77% | < 0.001 | 100.00% | < 0.001 |
| 4 | 87.10% | 0.002 | 100.00% | < 0.001 |
| 5 | 90.32% | < 0.001 | 100.00% | < 0.001 |
| 6 | 87.10% | 0.002 | 100.00% | < 0.001 |
| 7 | 100.00% | < 0.001 | 100.00% | < 0.001 |
| 8 | 93.55% | < 0.001 | 100.00% | < 0.001 |
| 9 | 77.42% | 0.065 | 100.00% | < 0.001 |
| 10 | 87.10% | 0.002 | 100.00% | < 0.001 |
| 11 | 90.32% | < 0.001 | 100.00% | < 0.001 |
| 12 | 87.10% | 0.002 | 100.00% | < 0.001 |
| 13 | 96.77% | < 0.001 | 100.00% | < 0.001 |
| 14 | 93.55% | < 0.001 | 100.00% | < 0.001 |
| 15 | 90.32% | < 0.001 | 100.00% | < 0.001 |
| 16 | 83.87% | 0.006 | 100.00% | < 0.001 |
| 17 | 80.65% | 0.026 | 100.00% | < 0.001 |
| 18 | 96.77% | < 0.001 | 100.00% | < 0.001 |
| 19 | 96.77% | < 0.001 | 100.00% | < 0.001 |

Exploratory endpoint values are described without adjustment for multiple comparisons and are reported as supportive observations rather than as evidence of efficacy on individual measures.

**Figure 1.**
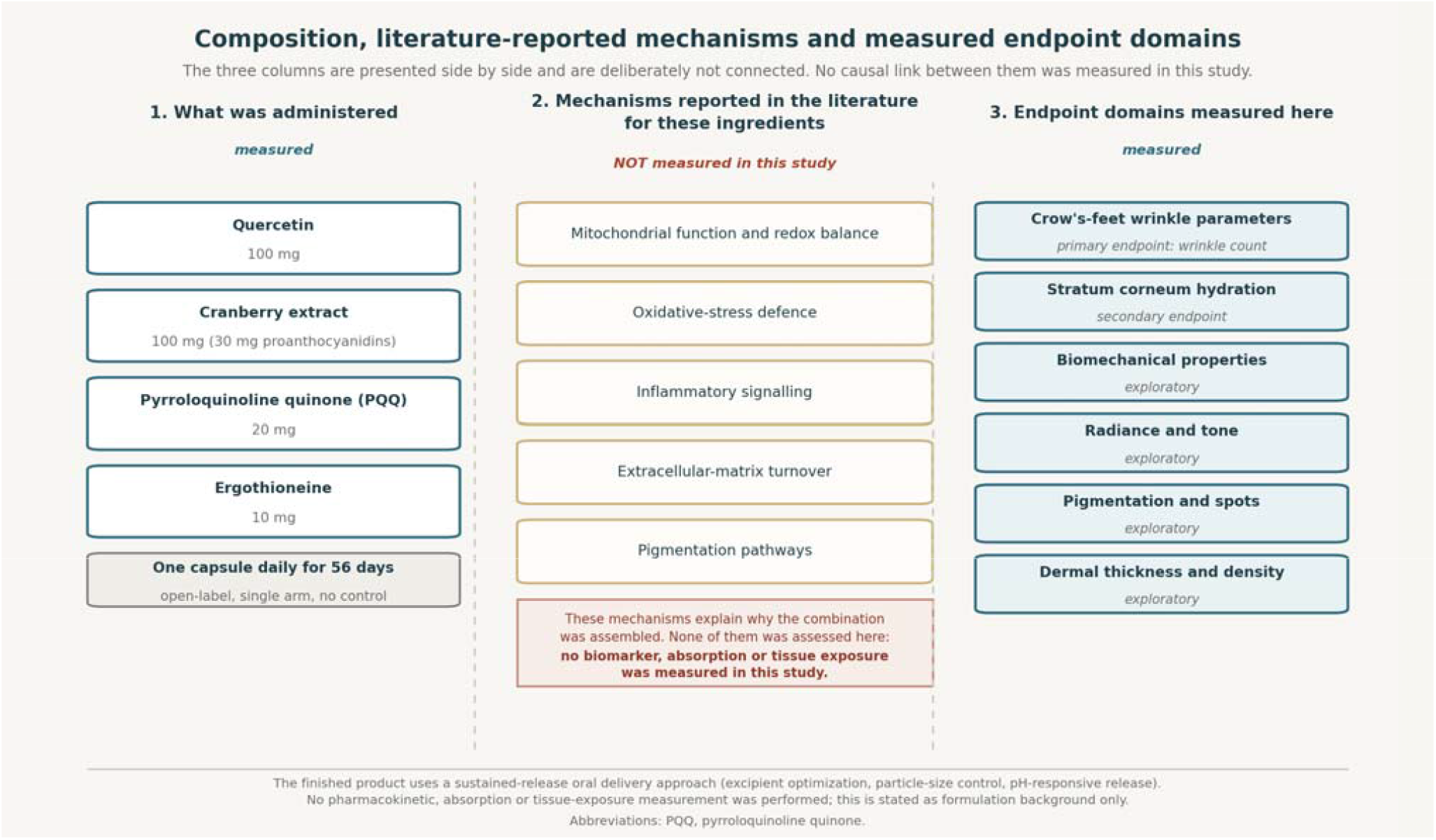
Composition, literature-reported mechanisms, and measured endpoint domains. The three columns list, respectively, what was administered, the mechanisms reported for these ingredients in the published literature, and the endpoint domains measured in this study. The columns are deliberately presented without connecting arrows: no link between an ingredient, a mechanism and an endpoint was measured or tested here, and none is asserted. Column 2 states why the combination was assembled and is not evidence about this product; no biomarker, absorption or tissue-exposure measurement was performed. Colour is used only to distinguish what was measured in this study (teal, columns 1 and 3) from what was not measured (gold, column 2).

Oral skin-support supplements have been studied as non-invasive adjuncts. A meta-analysis of randomized controlled trials reported that selected dietary supplements can improve skin hydration (Qian et al., 2022), and trials of collagen peptides, hyaluronan and multi-nutrient formulations have reported changes in hydration, elasticity, wrinkle parameters and dermal density using objective methods (Bolke et al., 2019; Göllner et al., 2017; Kim et al., 2022; Oe et al., 2021). Efficacy depends on ingredient selection, formulation, dosing and population; finished products therefore require dedicated clinical evaluation.

NatureU® Mind Care BeautyU Caps is a multi-ingredient oral beauty supplement. Each capsule contains quercetin 100 mg, cranberry extract 100 mg (equivalent to 20,000 mg cranberry raw material; 30 mg proanthocyanidins), pyrroloquinoline quinone (PQQ) 20 mg, and ergothioneine 10 mg. Published mechanistic literature offers a biological rationale only: PQQ has been associated with mitochondrial biogenesis via CREB phosphorylation and PGC-1α expression and with suppression of UVB-induced caspase-1 release in keratinocytes (Wong et al., 2009; Gruber and Holtz, 2022); quercetin has reported antioxidant, anti-inflammatory and photoprotective activities targeting JAK2/PKCδ and SIRT1 pathways (Shin et al., 2019; Li et al., 2022; Długosz et al., 2024); ergothioneine is reported to inhibit AP-1 and activate Nrf2 in UVA-irradiated dermal fibroblasts (Yang et al., 2020; Halliwell and Cheah, 2021); cranberry-derived proanthocyanidins are reported to modulate oxidative stress and pigmentation-related pathways (Xian et al., 2018; Pereira et al., 2021).

The finished product uses a sustained-release oral delivery approach, referred to by the sponsor as MASTER, based on excipient optimization, particle-size control and pH-responsive release. Delivery strategies of this type are discussed in the food-grade literature as means of improving the stability, solubility and intestinal release of polyphenols (Hu et al., 2016; Yang et al., 2020b; Mishra et al., 2022; Jiao et al., 2022). No pharmacokinetic, absorption or tissue-exposure measurement was performed in this study. This paragraph is therefore formulation background only; it forms no part of the study hypotheses or endpoints, and no claim about delivery performance is made or tested here. One author is a named inventor on a patent covering this delivery technology, which is held by the sponsor (see Conflicts of interest).

Figure 1 summarizes what was administered, the mechanisms reported for these ingredients in the published literature, and the endpoint domains measured in this study. The three columns are deliberately presented side by side and are not joined: no link between an ingredient, a mechanism and an endpoint was measured or tested here, and none is asserted.

The clinical performance of a finished multi-ingredient product cannot be inferred from ingredient literature alone. This 56-day exploratory single-arm study was designed to evaluate within-participant changes in PRIMOS CR crow’s-feet wrinkle count (primary endpoint), Corneometer-measured stratum corneum hydration (secondary endpoint), and additional exploratory skin-aging parameters in adult women, and to generate hypotheses for future randomized placebo-controlled studies. Reporting follows the TREND statement and the TIDieR checklist.

## 2. Materials and Methods

### 2.1 Study design

This was a single-center, open-label, single-arm exploratory clinical study conducted over 56 days, with assessments at baseline (D0), day 28 (D28) and day 56 (D56). The study was conducted by Centre Testing International Pinbiao (Shanghai) Co., Ltd., a contract testing organization, under commission from the sponsor, using standardized procedures for participant preparation, environmental control, instrument-based measurement, questionnaire collection and statistical analysis. Instrumental efficacy testing followed the published Chinese group standards for oral beauty products: T/CIET 411–2024 (moisturizing efficacy), T/CIET 415–2024 (anti-wrinkle efficacy) and T/CIET 406–2024 (whitening efficacy), together with the testing laboratory’s standard operating procedures. Reporting follows the TREND statement and the TIDieR checklist. The design, endpoint hierarchy and participant disposition are summarized in Table 1.

### 2.2 Participants

Healthy female volunteers aged 36–56 years were recruited. In total, 56 candidates were screened, 33 were enrolled and 31 completed the full protocol and were included in the completer analysis set.

Inclusion criteria were healthy women aged 36–56 years; cheek stratum corneum hydration below 60 C.U.; visible crow’s-feet wrinkles; facial laxity or reduced radiance; at least one visible facial spot meeting prespecified size and individual typology angle (ITA°) criteria; willingness to maintain regular daily habits throughout the study; and written informed consent. Participants further undertook, for the duration of the study, not to use any cosmetic, medication or health supplement capable of influencing the results; not to use any product with efficacy claims comparable to those of the study product; not to use any cosmetic or personal-care product other than the study product; and not to undergo any aesthetic-medicine procedure. These were undertakings given at enrolment rather than variables measured during the study; the distinction is addressed in the Limitations.

Exclusion criteria were skin conditions that could interfere with assessment; highly allergic constitution; pregnancy, lactation or planned pregnancy; severe systemic illness; serious psychiatric or endocrine disorders; oral contraceptive use; recent participation in other clinical studies; recent systemic or topical use of products that could influence skin outcomes; and inability to comply with study procedures.

Participant age is reported only as the eligibility range. No individual ages, dates of assessment, photographs or other potentially identifying details are reported here or in the figures.

### 2.3 Investigational product and intervention

The investigational product was NatureU® Mind Care BeautyU Caps, a hard-shell capsule oral supplement supplied by the sponsor. Each capsule contained quercetin 100 mg, cranberry extract 100 mg (equivalent to 20,000 mg cranberry raw material; standardized to 30 mg proanthocyanidins), PQQ 20 mg and ergothioneine 10 mg.

Before dispensing, the testing laboratory removed all brand and manufacturer identification from the product packaging, so that participants received the capsules in unlabelled form. This does not constitute blinding in the controlled-trial sense, since there was no comparator and participants knew they were taking an active supplement, but it removes brand identity as a source of expectancy. Participants took one capsule orally once daily for 56 consecutive days. Compliance was verified by product return and weighing.

### 2.4 Procedures and measurements

At baseline, eligible participants provided written informed consent and received instructions on daily intake and diary completion. At each visit, participants cleansed and dried the face and rested for 30 minutes in a controlled environment (21 ± 1 °C; relative humidity 50 ± 10%) before measurement. Measurements were taken at the same anatomical sites at each timepoint. The assessment battery comprised:

- **PRIMOS CR** (Canfield Scientific, Parsippany, NJ, USA; analysis software Primos 5.8 E) — crow’s-feet wrinkle count, length, volume, depth, area and area ratio, by three-dimensional fringe-projection profilometry. The segmentation threshold used to distinguish a wrinkle from background skin microrelief, the definition and area of the region of interest, and whether images from the three timepoints were spatially registered before analysis were requested from the contract testing organization but were not provided; these parameters are therefore not available for reporting. The implications are addressed in Section 4.
- **Corneometer CM 825** (Courage + Khazaka electronic GmbH, Cologne, Germany) — cheek stratum corneum hydration (C.U.).
- **Cutometer MPA580** (Courage + Khazaka) — gross elasticity R2, together with the derived ratio F3/F4 and the parameter F4 as implemented by the contract testing laboratory. R2 is a standard Cutometer output; F3/F4 and F4 are laboratory-specific derived parameters rather than standard Cutometer outputs and were recorded as exploratory only.
- **Glossymeter** (Courage + Khazaka) and **VISIA CR** (Canfield Scientific) — gloss and radiance.
- **Colorimeter CL400** (Courage + Khazaka) and **VISIA CR** — ITA°.
- **Mexameter MX18** (Courage + Khazaka) — melanin index.
- **VISIA CR** — spot area ratio, spot optical density, pore score and pore feature count.
- **DermaScan** (Cortex Technology, Hadsund, Denmark) — dermal thickness and density.
- **Structured self-assessment** at D28 and D56 (items 1–19, five-point agreement scale; item 20, self-reported adverse reactions; items 21–25, perceived onset). The full wording of all 25 items is given in Appendix Table A2.

### 2.5 Endpoint hierarchy

Primary endpoint: within-participant change from baseline at D56 in PRIMOS CR crow’s-feet wrinkle count. Secondary endpoint: within-participant change from baseline at D56 in Corneometer stratum corneum hydration. All remaining instrumental parameters and participant-reported outcomes were exploratory. D28 measurements were treated as supportive interim observations.

### 2.6 Statistical analysis

The prespecified analysis set was a completer analysis (n = 31). No imputation was performed. No formal power calculation was performed because the study was exploratory.

Continuous variables are summarized as mean ± standard deviation. Within-participant changes were assessed by paired Student’s t-test when normality (Shapiro–Wilk) was supported and by the Wilcoxon signed-rank test otherwise. Change rate (%) was computed as ((post-intervention value − baseline value) / baseline value) × 100 from unrounded means; the original testing report computed the same quantity from means rounded to one decimal place, which differs in the second decimal place for some parameters. P values are nominal; no multiplicity adjustment was applied.

Participant-reported recognition rates were assessed against a prespecified benchmark of 60% agreement using a two-sided exact binomial test with n = 31, as specified in the contract testing protocol. The D28 P values given in the original testing report were not reproducible under that specification: five items sharing an identical numerator of 27 of 31 (87.10%) were reported with five different P values (0.006, 0.016, 0.037, 0.076 and 0.350), which no single test can produce, and the ranking of P values was inverted relative to the recognition rates. All questionnaire P values have therefore been recomputed by the authors under the specification above and are the values given in Table 5. This issue has been reported to the contract testing organization.

Statistical analyses in the original testing report were performed by the contract testing organization using SPSS (IBM Corp., Armonk, NY, USA); the software version is not stated in the report and was requested from the contract testing organization but was not provided. The recomputed values and all individual-participant analyses reported in this version — proportional-change analysis, correlation analyses, the bounding analysis for withdrawals and the quantification of regression to the mean — were computed by the authors in Python 3.10 (Python Software Foundation, Wilmington, DE, USA) using SciPy 1.15.3 and NumPy 2.2.6, on the individual-participant dataset received from the contract testing organization. All summary statistics in the testing report were reproduced from that dataset before any new analysis was undertaken.

## 3. Results

### 3.1 Participant disposition and tolerability

Of 33 enrolled participants, 31 (93.9%) completed the protocol and were included in the completer analysis set. Two withdrew, both for reasons unrelated to the study product: one was lost to follow-up at the D56 visit after contracting influenza A, and one withdrew on day 4 because of illness. The contract testing organization reports that neither has evaluable data, so a full-analysis-set analysis of all 33 is not possible. Design, disposition and tolerability are summarized in Table 1.

As a bounding analysis, assuming zero change for both withdrawals at the observed baseline mean and recomputing over n = 33, the reduction in wrinkle count moves from −46.73% to −43.89% and the increase in hydration from +58.57% to +55.02%, both remaining significant (Wilcoxon P = 1.2 × 10□□ for each); the change in wrinkle depth remains non-significant. The conclusions are not altered by the two withdrawals.

No adverse reactions were reported by any of the 31 evaluable participants.

### 3.2 Primary endpoint — crow’s-feet wrinkle count

PRIMOS CR crow’s-feet wrinkle count decreased from 965.3 ± 333.7 at D0 to 518.2 ± 155.6 at D28 and 514.2 ± 171.0 at D56 (−46.32% at D28 and −46.73% at D56; Table 2). Differences from baseline were not normally distributed (Shapiro–Wilk P = 0.002), so significance is reported from the Wilcoxon signed-rank test (P = 1.2 × 10 □□).

Three features of the individual-level data require reporting; all three are visible in Figure 3.

First, the count decreased in all 31 participants, without exception (Figure 3a).

Second, the decrease was almost exactly proportional across the cohort. The ratio of D56 to D0 count had a mean of 0.535 with a standard deviation of 0.041 — a coefficient of variation of 7.7% — across baseline values spanning a 5.9-fold range (360 to 2116), and this ratio was not correlated with the baseline value (r = −0.15, P = 0.42). Linear regression of D56 on D0 gave D56 = 0.499 × D0 + 33, with R^2^ = 0.948 (Figure 3d). For comparison, the coefficient of variation of the corresponding D56/D0 ratio was 20.3% for hydration and 16.3% for wrinkle depth, so the primary endpoint showed less between-participant dispersion than either.

Third, the change was complete by D28 and did not progress thereafter: the ratio of D56 to D28 count was 0.990 ± 0.096, and the count increased slightly between D28 and D56 in 11 of 31 participants (Figure 3a).

### 3.3 Secondary endpoint — stratum corneum hydration

Hydration increased from 44.3 ± 7.8 C.U. at D0 to 73.1 ± 10.3 at D28 and 70.3 ± 9.9 at D56 (+64.77% at D28 and +58.57% at D56; paired t-test P = 4.6 × 10□^1^□; Wilcoxon P = 1.2 × 10□□; differences normally distributed, Shapiro–Wilk P = 0.61; Table 2). Hydration increased in all 31 participants, but with substantially greater between-participant dispersion than the primary endpoint (D56/D0 ratio 1.626 ± 0.330; Figure 3b). Four of 31 participants remained below the 60 C.U. entry threshold at D56.

### 3.4 Exploratory endpoints

Other PRIMOS wrinkle parameters showed reductions in length (−23.68%), volume (−21.29%), area (−17.13%) and area ratio (−17.06%) at D56 (Table 3). Wrinkle depth showed only a non-significant numerical decrease, from 42.26 ± 10.02 to 40.23 ± 9.93 (−4.81%; paired t-test P = 0.074, Wilcoxon P = 0.058); depth decreased in 19 of 31 participants, increased in 11 and was unchanged in one (Figure 3c). The individual change in wrinkle count was only weakly related to the individual change in wrinkle depth (r = +0.30, P = 0.096, computed on absolute changes) and was not related to the individual change in hydration (r = +0.23, P = 0.21, computed on D56/D0 ratios; on absolute changes r = −0.03, P = 0.87).

Non-wrinkle instrumental outcomes are given in Table 4. Biomechanical properties: Cutometer R2 increased by 19.05% and the F3/F4 ratio by 90.72%. The laboratory-specific firmness parameter F4 increased from 0.699 ± 0.451 to 1.792 ± 0.672 (+156.37%), the largest percentage change in the dataset; as set out in Section 4.3, this reflects the small magnitude of its baseline value rather than the largest biological effect observed.

Radiance and tone: Glossymeter gloss +28.96%, VISIA CR gloss +47.10%, Colorimeter ITA° +16.13%, VISIA CR ITA° +9.09%. Pigmentation and spots: Mexameter melanin index −14.79%, VISIA CR spot area ratio −15.52%, spot optical density −4.60%. Pores: VISIA CR pore feature count −9.31%. Dermal structure: DermaScan thickness +16.36% and density +5.36%. Figure 2 provides a visual overview of the within-participant percentage changes at D56 across all instrumental outcomes.

**Figure 2.**
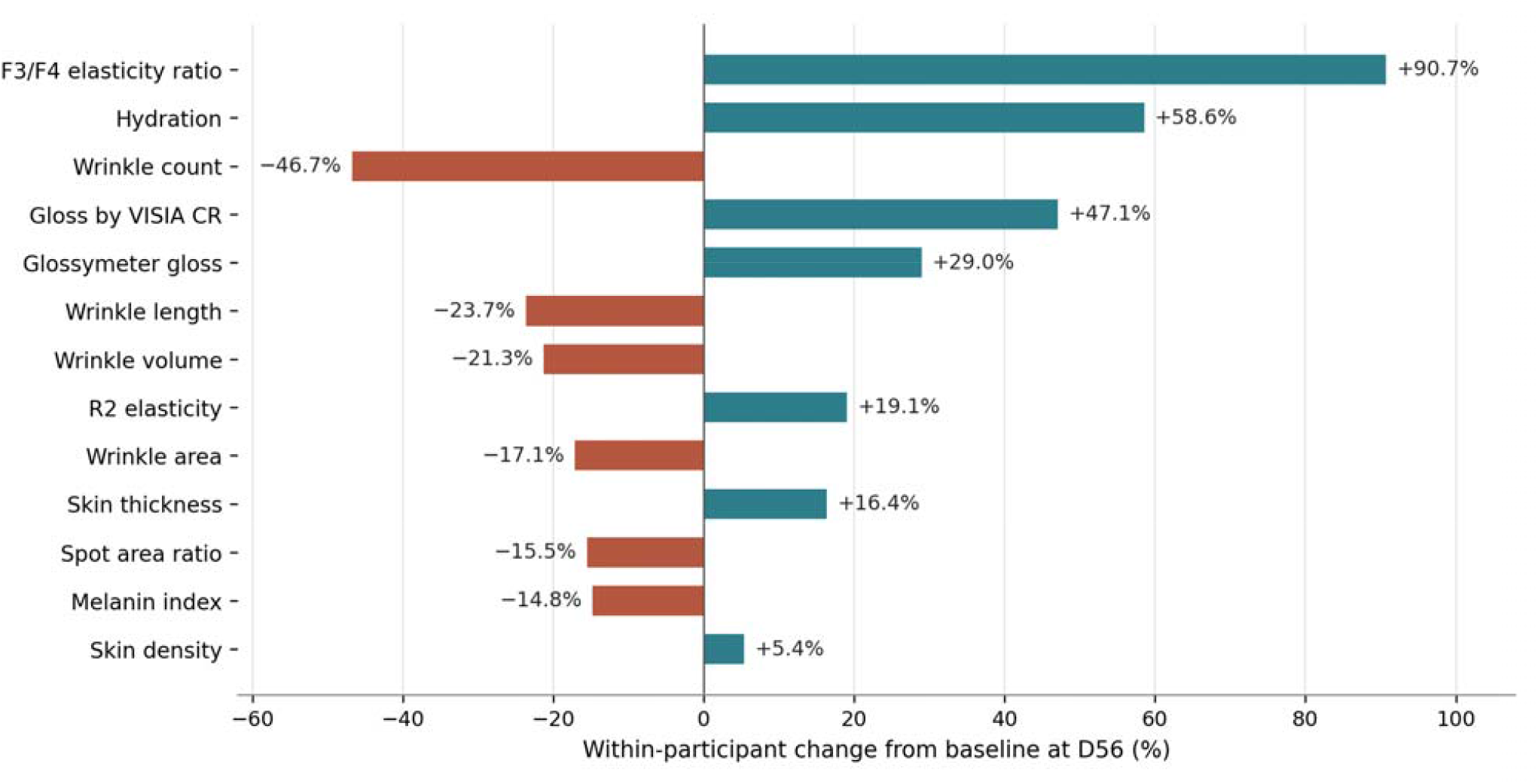
Within-participant percentage changes in key instrumental skin outcomes at day 56 (n = 31 completers). Positive bars represent increases in hydration, biomechanical, gloss or dermal-structure measures; negative bars represent reductions in pigment-, spot- or wrinkle-burden measures. Provided as a visual overview only; full numerical values are given in Tables 2–4. The hydration bar is shown as +58.6%, the value computed from unrounded means.

Participant-reported outcomes: under the recomputed binomial values, all items except item 9 at D28 (77.42%, P = 0.065) exceeded the prespecified 60% benchmark at both timepoints (Table 5). The distribution of perceived onset is given in Appendix Table A1. Participant-reported outcomes are de-emphasized throughout because self-assessment in an unblinded study is particularly susceptible to expectancy.

Exploratory endpoint values are described without adjustment for multiple comparisons and are reported as supportive observations rather than as evidence of efficacy on individual measures.

## 4. Discussion

This study was designed to generate signals, not to demonstrate efficacy. The interpretation below differs materially from that of version 1 because the individual-participant data were not available when version 1 was written.

### 4.1 The primary endpoint

The choice of wrinkle count as the prespecified primary endpoint requires direct comment. Count and depth are not interchangeable quantities. A count-based parameter enumerates discrete features that exceed a segmentation threshold, and therefore responds to any change that moves shallow features across that threshold, including changes in surface hydration and light scattering; a depth-based parameter instead quantifies the vertical dimension of features that are already resolved. The two need not move together, and in this dataset they did not. We read this dissociation as indicating a change in the visibility of fine surface texture rather than structural remodelling of established wrinkles. A depth-or volume-based parameter would have been the more conservative choice, and we would select one in a confirmatory trial. The primary endpoint was nevertheless fixed before analysis and is reported as prespecified; we have not resolved the problem by substituting a different primary endpoint after seeing the data.

The individual-participant data raise a further question that we consider more important than the choice of endpoint. The reduction in wrinkle count has three features that are difficult to reconcile with a biological effect: it occurred in every participant without exception; it was almost exactly proportional across a 5.9-fold range of baseline values, with the lowest between-participant dispersion of any endpoint measured here and no correlation with baseline severity; and it was complete at the first follow-up visit, increasing slightly in 11 of 31 participants between D28 and D56. A biological response to an oral supplement would not ordinarily produce a near-constant proportional change that is independent of baseline burden, complete within four weeks with no subsequent progression, and uncorrelated with the concurrent change in either wrinkle depth or hydration.

This pattern is at least as consistent with a systematic difference between the baseline imaging session and the two follow-up sessions — a difference in segmentation threshold, region-of-interest placement, image registration or operator — as it is with a treatment effect. We requested these acquisition and analysis parameters from the contract testing organization; the device and software version were supplied but the analysis parameters were not, and we are therefore unable to exclude a methodological explanation. **We do not claim that the reduction in crow’s-feet wrinkle count observed here represents a biological anti-wrinkle effect, and we recommend that the primary endpoint of this study be treated as uninterpretable until the analysis parameters are documented**.

### 4.2 The hydration endpoint and regression to the mean

The hydration result does not share those features: it varied substantially between participants, and four participants remained below the entry threshold at D56.

The contribution of regression to the mean can be bounded rather than merely acknowledged. Eligibility required baseline hydration below 60 C.U.; the cohort baseline mean was 44.33 C.U. and the rise to D56 was 25.96 C.U. Taking the expected regression component as (1 − ρ)(μ − 44.33), where ρ is the test–retest reliability of the measurement and μ the mean of the unselected source population: a conservative reliability of 0.80 with a source-population mean of 55 C.U. gives 2.13 C.U., or 8.2% of the observed rise; the most permissive assumptions we can construct (ρ = 0.70, source-population mean equal to the observed follow-up value of 70.3 C.U.) give 7.79 C.U., or 30.0%. More directly, for regression to the mean alone to account for the entire rise, the mean cheek hydration of the unselected population would have to be at least 70.3 C.U., which would make an entry criterion of below 60 C.U. meaningless as a selection filter.

Regression to the mean is therefore a real but partial contributor and cannot plausibly explain the hydration result in full. This does not establish a treatment effect: seasonal drift, behavioural change and expectancy remain uncontrolled in a single-arm design.

### 4.3 The F4 parameter

One exploratory value merits specific comment because its magnitude is conspicuous. F4 increased by 156.37% at D56, more than any other measure reported here. This reflects the arithmetic of the parameter rather than the size of the underlying change: F4 had a baseline mean of 0.699 with a standard deviation of 0.451, a coefficient of variation of approximately 65% and the highest relative dispersion in the dataset, and the absolute change was 1.09 units. When a derived parameter has a small mean lying close to the lower bound of its range, a modest absolute shift yields a large percentage. Percentage change is not a suitable basis for ranking effects across parameters measured on different scales.

### 4.4 Attribution to individual components

A single fixed combination was administered without factorial, dose-ranging or single-ingredient arms. The design cannot attribute any observed change to quercetin, cranberry-derived proanthocyanidins, PQQ or ergothioneine individually, nor establish whether the components act additively, synergistically or not at all in combination. The ingredient literature cited here explains only why the combination was assembled; it provides no evidence about the contribution of any single component. Component-dropout and dose-ranging designs would be required before any attribution could be made.

Because absorption, pharmacokinetics, tissue exposure, OCTN1-mediated transport (Arakawa et al., 2017; Ben Mariem et al., 2024) and biomarker changes were not measured here, mechanistic statements remain biological rationale rather than demonstrated clinical mechanism.

### 4.5 Limitations

The study was open-label and single-arm without a placebo control, so causal inference is not possible. The sample size was modest and the analysis was restricted to 31 completers without imputation. No mechanistic biomarkers were measured. Exploratory endpoints were not adjusted for multiple comparisons. The cohort was recruited at a single centre within a narrow age window and a single ethnicity-relevant subset, limiting generalizability. The study was registered only after data collection had been completed; retrospective registration means that the endpoints and analysis plan cannot be independently verified against a public record predating the study.

Several items of methodological information were requested from the contract testing organization and were not provided: the PRIMOS segmentation threshold, region-of-interest definition and image-registration procedure; baseline participant characteristics beyond age, including skin phototype, body mass index, smoking status, sun-exposure habits and baseline skincare use; the composition and constitution of the ethics review committee; which behavioural variables were actively recorded as opposed to being covered by participant undertakings; and the custody of and access to the raw data, the authorship of the statistical analysis plan and whether an independent statistician reviewed the final analysis. We report these gaps rather than infer answers to them.

A randomized, double-blind, placebo-controlled trial with a prespecified depth- or volume-based primary endpoint, documented image-analysis parameters, larger sample size, baseline stratification, longer follow-up and mechanistic biomarkers is required to confirm or refute the present exploratory signals.

## 5. Conclusion

In this 56-day open-label, single-arm exploratory study in adult women, daily intake of NatureU® Mind Care BeautyU Caps was associated with a within-participant increase in stratum corneum hydration and with directionally congruent changes across several exploratory skin-aging endpoints, with no reported adverse reactions. A reduction in crow’s-feet wrinkle count was also recorded, but its uniformity across participants, its independence of baseline severity, its completion at the first follow-up visit and the absence of the analysis parameters needed to reproduce it mean that we do not interpret it as evidence of a biological anti-wrinkle effect. This conclusion supersedes that of version 1. Findings are hypothesis-generating; randomized, placebo-controlled trials are required.

### Ethics statement

The study protocol, participant information sheet and informed consent form were reviewed and approved on 3 January 2025 (reference A225000034310100101C) by the ethics committee of Centre Testing International Pinbiao (Shanghai) Co., Ltd., Shanghai, China.

This is the internal ethics committee of the contract testing organization that conducted the study under commission from, and for payment by, the sponsor. **It is not an ethics committee independent of the organization that performed and invoiced the work**. Under the Chinese Measures for Ethical Review of Life Science and Medical Research Involving Human Subjects, institutions conducting such research establish their own ethics review committees, so an institutional committee of this kind is the expected arrangement rather than an irregular one; readers should nevertheless be aware that approval and conduct sat within the same organization. The committee’s composition, and confirmation that personnel involved in conducting the study were recused from the review, were requested from the contract testing organization and were not provided.

The study was conducted in accordance with the principles of the Declaration of Helsinki. All participants provided written informed consent before any study-related procedure, including consent for the publication of anonymized study data. The informed consent form (version V1.0, dated 20 December 2024) predates the start of data collection on 11 January 2025. No individual ages, assessment dates, photographs or participant identifiers are reported in this manuscript.

### Trial registration

ClinicalTrials.gov identifier **NCT07571629. This study was registered retrospectively**. Data collection ran from 11 January to 8 March 2025; the record was first submitted on 30 April 2026 and first posted on 6 May 2026 — approximately 14 months after completion of data collection. The endpoints and analysis plan were prespecified in the contract testing protocol but were not publicly preregistered before the study began. Version 1 of this preprint stated in its body text that the study “was not prospectively registered” while supplying a registration identifier in the submission metadata without this explanation; the present statement corrects that inconsistency.

## Data availability

The individual-participant dataset for crow’s-feet wrinkle count, wrinkle depth and stratum corneum hydration at D0, D28 and D56 underlies all analyses newly reported in this version, and the summary statistics derived from it are reported in full in Tables 2–5 and Figure 3. The clinical testing report and additional de-identified study documentation are available from the corresponding author on reasonable request, subject to confidentiality, participant privacy and applicable regulatory requirements.

**Figure 3.**
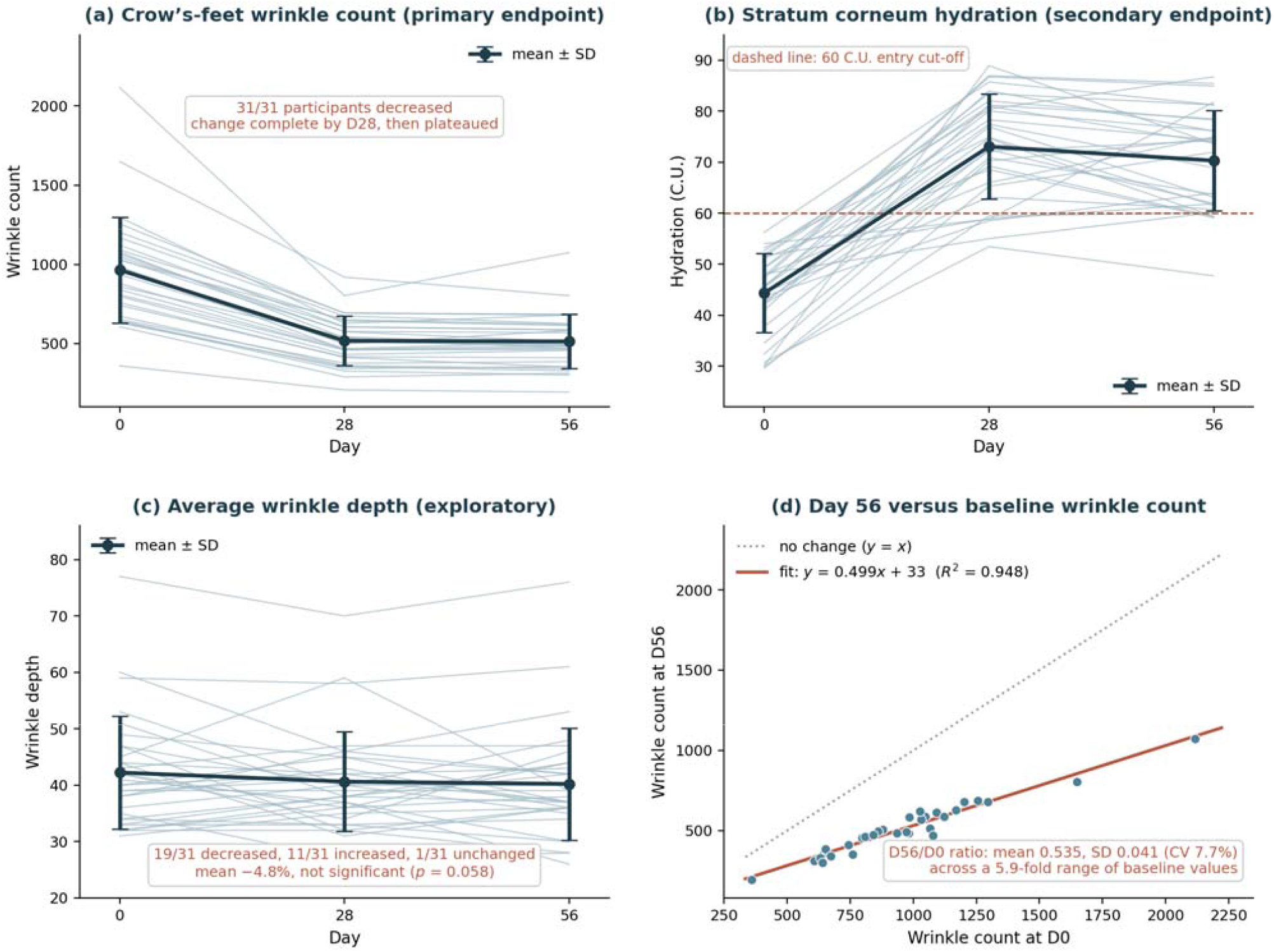
Individual-participant trajectories over 56 days in the 31 completers. (a) Crow’s-feet wrinkle count, the prespecified primary endpoint. (b) Stratum corneum hydration, the prespecified secondary endpoint; the dashed line marks the 60 C.U. entry criterion. (c) Average wrinkle depth. (d) Day 56 versus baseline wrinkle count, with the line of identity and the fitted regression. Faint lines and points are individual participants; the heavy line shows mean ± standard deviation. Panels (a) and (d) show that the count fell in every participant by a near-constant proportion irrespective of baseline value, and panel (a) that the change was complete by day 28. This pattern is discussed in Section 4.1. No participant identifiers are shown.

## Acknowledgements

The authors thank the study participants, and colleagues at Centre Testing International Pinbiao (Shanghai) Co., Ltd. for conducting the study procedures and instrumental assessments.

During the preparation of this manuscript the authors used Claude Opus 5 (Anthropic) to assist with reference formatting and verification against source records, language editing, statistical recomputation from the individual-participant dataset, and figure preparation; Perplexity AI was used to assist with literature searching during preparation of an earlier draft. No generative AI tool was used to generate, analyse in place of the authors, alter or interpret any study data, images or results. The authors reviewed and edited all AI-assisted output and take full responsibility for the content and integrity of this publication.

## Appendix

**Appendix Table A1.** Participant-reported perceived onset of benefit at D56 (questionnaire items 21–25; n = 31 completers). Values are the percentage of participants selecting each interval.

| Item | ≤ 7 days | 8–14 days | 15–28 days | 29–56 days | No perceived effect by D56 |
| --- | --- | --- | --- | --- | --- |
| 21 — skin tone / pigmented spots | 6.45% | 16.13% | 48.39% | 29.03% | 0.00% |
| 22 — wrinkles | 3.23% | 19.35% | 32.26% | 45.16% | 0.00% |
| 23 — hydration | 22.58% | 29.03% | 38.71% | 9.68% | 0.00% |
| 24 — elasticity and firmness | 9.68% | 22.58% | 29.03% | 38.71% | 0.00% |
| 25 — refinement and radiance | 3.23% | 19.35% | 45.16% | 32.26% | 0.00% |

**Appendix Table A2.**
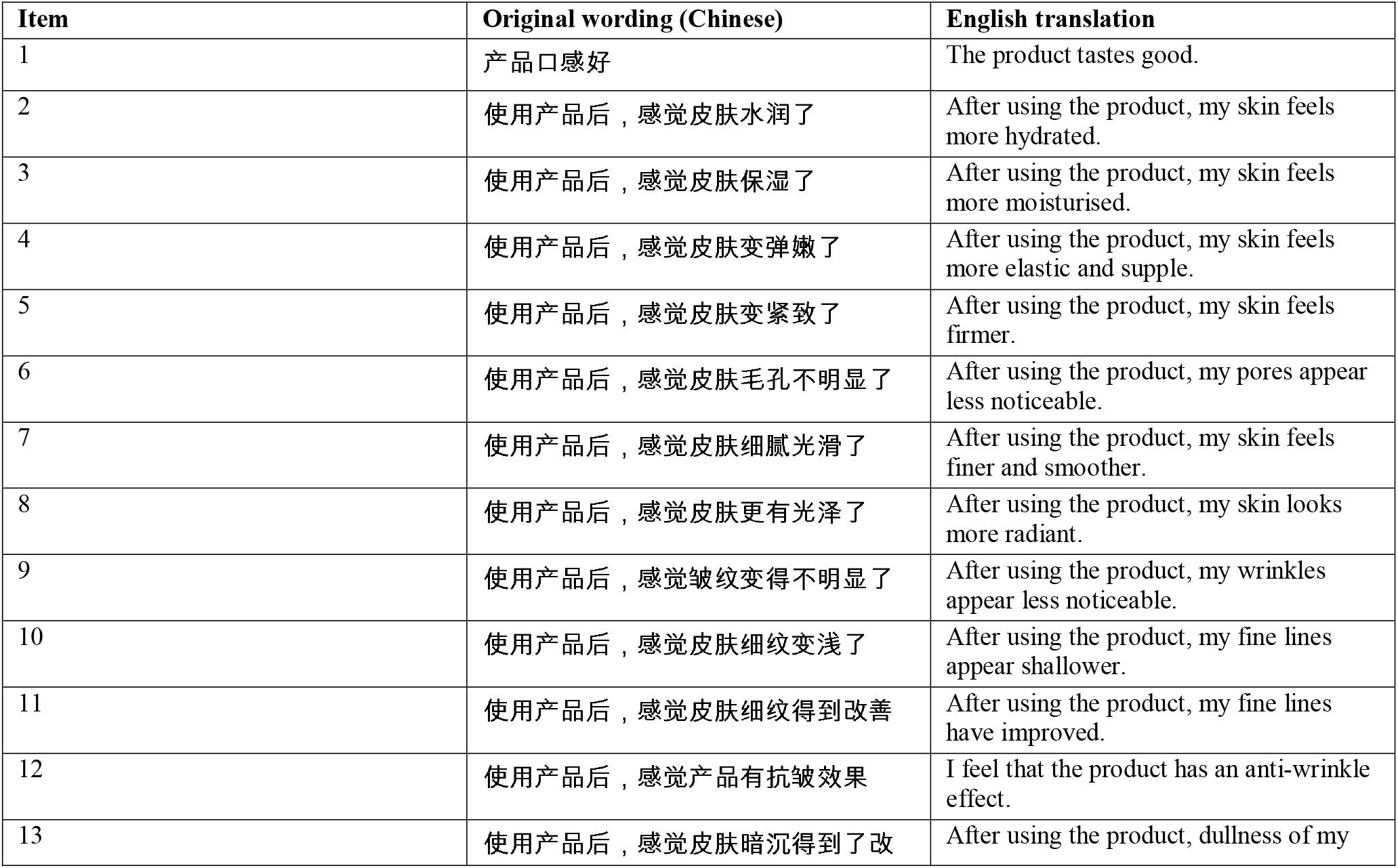
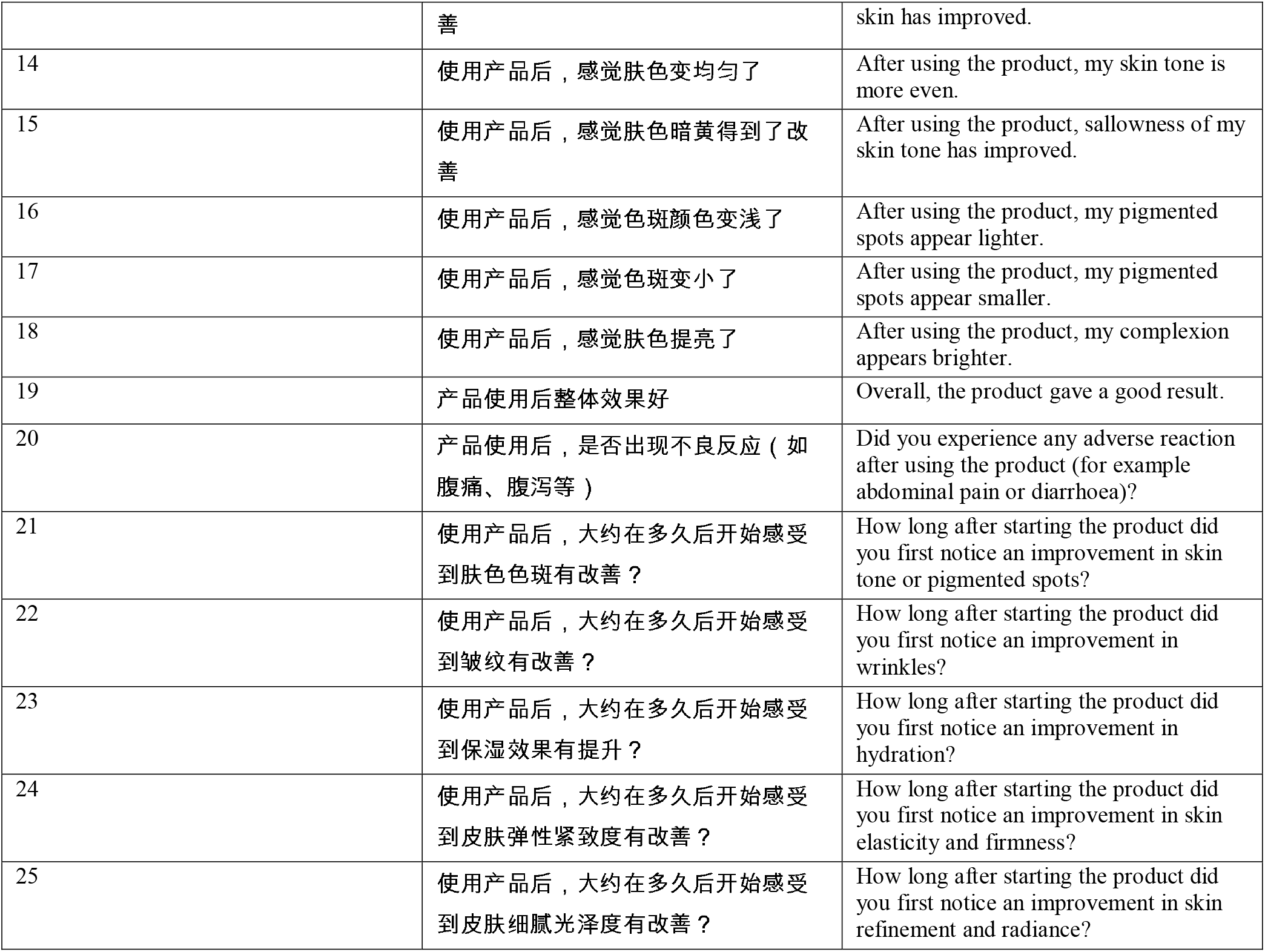
Full wording of the participant self-assessment questionnaire. Items 1–19 were scored on a five-point agreement scale (1 = strongly disagree, 2 = disagree, 3 = neither agree nor disagree, 4 = agree, 5 = strongly agree); the recognition rate reported in Table 5 is the proportion of participants selecting an agreement-level response (4 or 5). Item 20 was a yes/no safety item. Items 21–25 used the intervals shown in Appendix Table A1. The questionnaire was administered in Chinese; the English translations were prepared for publication and were not used during data collection.

| Item | Original wording (Chinese) | English translation |
| --- | --- | --- |
| 1 | 产品口感好 | The product tastes good. |
| 2 | 使用产品后，感觉皮肤水润了 | After using the product, my skin feels more hydrated. |
| 3 | 使用产品后，感觉皮肤保湿了 | After using the product, my skin feels more moisturised. |
| 4 | 使用产品后，感觉皮肤变弹嫩了 | After using the product, my skin feels more elastic and supple. |
| 5 | 使用产品后，感觉皮肤变紧致了 | After using the product, my skin feels firmer. |
| 6 | 使用产品后，感觉皮肤毛孔不明显了 | After using the product, my pores appear less noticeable. |
| 7 | 使用产品后，感觉皮肤细腻光滑了 | After using the product, my skin feels finer and smoother. |
| 8 | 使用产品后，感觉皮肤更有光泽了 | After using the product, my skin looks more radiant. |
| 9 | 使用产品后，感觉皱纹变得不明显了 | After using the product, my wrinkles appear less noticeable. |
| 10 | 使用产品后，感觉皮肤细纹变浅了 | After using the product, my fine lines appear shallower. |
| 11 | 使用产品后，感觉皮肤细纹得到改善 | After using the product, my fine lines have improved. |
| 12 | 使用产品后，感觉产品有抗皱效果 | I feel that the product has an anti-wrinkle effect. |
| 13 | 使用产品后，感觉皮肤暗沉得到了改 | After using the product, dullness of my |
|  | 善 | skin has improved. |
| 14 | 使用产品后，感觉肤色变均匀了 | After using the product, my skin tone is more even. |
| 15 | 使用产品后，感觉肤色暗黄得到了改善 | After using the product, sallowness of my skin tone has improved. |
| 16 | 使用产品后，感觉色斑颜色变浅了 | After using the product, my pigmented spots appear lighter. |
| 17 | 使用产品后，感觉色斑变小了 | After using the product, my pigmented spots appear smaller. |
| 18 | 使用产品后，感觉肤色提亮了 | After using the product, my complexion appears brighter. |
| 19 | 产品使用后整体效果好 | Overall, the product gave a good result. |
| 20 | 产品使用后，是否出现不良反应（如腹痛、腹泻等） | Did you experience any adverse reaction after using the product (for example abdominal pain or diarrhoea)? |
| 21 | 使用产品后，大约在多久后开始感受到肤色色斑有改善？ | How long after starting the product did you first notice an improvement in skin tone or pigmented spots? |
| 22 | 使用产品后，大约在多久后开始感受到皱纹有改善？ | How long after starting the product did you first notice an improvement in wrinkles? |
| 23 | 使用产品后，大约在多久后开始感受到保湿效果有提升？ | How long after starting the product did you first notice an improvement in hydration? |
| 24 | 使用产品后，大约在多久后开始感受到皮肤弹性紧致度有改善？ | How long after starting the product did you first notice an improvement in skin elasticity and firmness? |
| 25 | 使用产品后，大约在多久后开始感受到皮肤细腻光泽度有改善？ | How long after starting the product did you first notice an improvement in skin refinement and radiance? |

